# Dynamic pressure ulcer risk predictions for hospitalized patients: development and validation of a machine learning model with an expert group of nurses

**DOI:** 10.1101/2024.12.16.24319086

**Authors:** Steffen Greup, Denise Spoon, Annemarie de Vroed, Ben Werkhoven, Enrico Timmerman, Erwin Ista, Monique van Dijk

## Abstract

**Background:** Pressure ulcers (PU) are a source of harm and discomfort in hospitalized patients. To identify patients at risk for developing PU nurses use validated risk assessment instruments which have insufficient predictive power, also leading to increased nursing workload. Machine learning on electronic health records (EHR) has the potential to provide better risk predictions while also reducing nursing workload.

**Objective:** To develop and validate a dynamic prediction model for daily PU risk predictions together with an expert group of nurses, while considering factors important to successful clinical implementation like explainability and robustness during the modelling process.

**Methods:** All admissions to general wards in a single-center tertiary university hospital in the Netherlands were included. A retrospective dataset with candidate predictors was collected from the EHR and split into a training set (December 2021 – November 2022, N=19931) and a validation set (January 2023 – June 2023, N=11387). Separate models were developed for the first 72 hours of admission, and >72 hours. The PU outcome was identified from both structured and free text registration. Predictor definition, selection and modelling choices were discussed with an expert group of nurses. Several candidate machine learning models were compared using cross-validation on the training set. The same final model and predictor set were selected for both ≤72h and >72h, and predictive performance on the validation set was compared to the Waterlow score.

**Results:** The difference in performance across candidate models was small on the training set. Logistic regression with an L2 penalty and a spline transformation applied to a limited set of predictors was chosen as the final model, and the candidate predictor set was reduced to a final predictor set. The AUROC for the final model was 0.790 (≤72h) and 0.795 (>72h) on the validation dataset. The model clearly outperformed the Waterlow score (0.816 vs. 0.702 (≤72h) and 0.800 vs. 0.677 (>72h)) for the subset of admissions with a registered Waterlow score. Expected remaining length of stay, age and ADL (activities of daily living) score were the predictors with the strongest contribution.

**Conclusions:** We developed and validated a dynamic prediction model for daily PU risk predictions, which outperformed the traditional Waterlow risk assessment score. During the development process special care was given to considerations for implementation and user acceptance. The model was named DRAAI (Decubitus Risk Alert based on AI), which means ‘turn’ in Dutch and is a common PU preventive measure.

## Background

Pressure ulcers (PU) are a source of harm and discomfort in hospitalized patients, and may lead to prolonged hospital stay, additional costs and higher workload for healthcare professionals (Demarré et al., 2015; Goodall et al., 2020). Causes of PU are often multifactorial, including patient-related risk factors, such as being underweight and immobile as well as surface-related factors such as suboptimal matrasses or seating cushions. Additionally, disease and treatment-related factors may contribute to the development of pressure ulcers, for instance diabetes, corticosteroid treatments or vasopressors (Coleman et al., 2014; Wang et al., 2023).

To identify patients at risk for developing PU nurses use validated standard risk assessment tools such as the Waterlow, Braden and Norton scales (Moore & Patton, 2019). However, research has shown their predictive properties to vary widely between studies (Pancorbo-Hidalgo et al., 2006; Park & Lee, 2016). Furthermore, due to changes in risk factors, the assessments should be performed regularly during a patient’s admission, leading to increased nursing workload. Therefore, several clinical prediction models have been developed, usually reporting higher predictive performance scores than those reported for standard risk assessment tools (Park & Lee, 2016; Zhou et al., 2023). However, developing a model intended for clinical implementation requires considering several other aspects than predictive performance alone. Explainability has the potential to increase user acceptance (Markus et al., 2021). However, explainability alone is not enough: if explanations do not conform with well-established knowledge, the model lacks credibility (Wang et al., 2018).

This can happen when the direction of the effect of the predictor is opposite to expert opinion, for example if a model would predict that PU risk decreases with increased surgery duration. The direction of effects in prediction models can change due to differences in included covariates and has also been shown to vary between healthcare settings and databases (Markus et al., 2022). Finally, the model should be able to sustain performance over time after clinical implementation, while clinical care evolves and EHR systems can change. Both the choice of predictors and differences between machine learning models can influence susceptibility to performance degradation over time (Jung & Shah, 2015; Nestor et al., 2019). For instance, due to a change in policy related to PU prevention or treatment, the role of different predictor variables may change over time.

The aim of this study was to develop and validate a dynamic prediction model for daily PU risk predictions together with an expert group of nurses, while considering factors important to successful clinical implementation like explainability, credibility and robustness over time during the development process.

## Methods

### Study design and setting

This study was conducted using data from all admissions of adult patients who were hospitalized for at least 24 hours to general wards in the Erasmus University Medical Center, a single-center tertiary hospital in Rotterdam, the Netherlands, between December 2020 and June 2023. The intensive care, pediatric and psychiatric wards were excluded due to expected differences in risk factors and availability of candidate predictors. The reporting of the study is in accordance with the ‘transparent reporting of a multivariable prediction model for individual prognosis or diagnosis’ (TRIPOD) statement (Collins et al., 2015). In Table 1 the machine learning terminology used in this paper are defined.

**Table 1.** Machine learning terminology used in this article.

| Term | Explanation |
| --- | --- |
| **Outcome** | The outcome is also called the target or the dependent variable. |
| **Candidate predictors** | Predictors are also called independent variables, covariates, or features. In this study they are the patient-level variables which are used to predict the outcome. From the complete set of candidate predictors, a subset will be selected as the final predictor set. |
| **Candidate models** | A set of machine learning models (such as random forest or logistic regression) from which a final model will be selected. |
| **Training set** | Data used for model development and training. |
| **Test set** | Data used for model validation. |
| **Hyperparameter** | A parameter controlling the configuration of a machine learning model. For example, the number of trees in a random forest is a hyperparameter. To process of hyperparameter optimization is also called the tuning of a hyperparameter. |
| **K-fold cross-validation** | This procedure is used on the training set. The set is split into k subsets. A model is trained on all subsets but one holdout subset, and the predictive performance is calculated on that holdout subset. This process is repeated until all k subsets have been the holdout subset. Thus, k values for predictive performance result. If the process of splitting the data into subsets is repeated multiple times (with randomness in the sampling resulting in different subsets), it is called repeated k-fold cross-validation. When the outcome rate is held constant between folds, it is called stratified cross-validation. |
| **Discrimination** | In the case of binary prediction (such as in this study), discrimination is the ability of a machine learning model to correctly separate the patients into groups. In the case of perfect discrimination, the model will predict the highest risk for all patients with the outcome event. |
| **Calibration** | The correspondence between the predicted probabilities and the actual probabilities. For example, if for a group of patients the mean predicted probability = 0.1, this is compared to the proportion with the actual outcome event (which ideally should also be 0.1). |
| **Decision curve** | A method to evaluate prediction models, sought to overcome the limitations of traditional metrics, such as discrimination and calibration, which are not directly informative as to clinical value. The decision curve analysis calculates a clinical ‘net benefit’ of the prediction model compared to treating all or none of the patients. |
| **Temporal validation** | A validation method where the data is split into subsets based on calendar date. A model is trained on the subset earlier in time, and validated on the subset later in time. |
| **Winsorization** | A data preprocessing method for numerical predictors. Extreme values are limited to reduce the influence of outliers. |
| **Yeo-Johnson transformation** | A data preprocessing method for numerical predictors. It is used to transform data with a skewed distribution to a normal distribution. |
| **Spline transformation** | A data preprocessing method for numerical predictors. It is used to allow a non-linear relationship between a predictor and the outcome. The relationship between predictor and outcome is divided into several windows, which are defined by so-called knots. Within each window, the relationship between predictor and outcome is modelled by a polynomial. |
| **One-hot encoding** | A data preprocessing method for categorical predictors. It converts a single column into multiple columns, one for each level of the categorical predictor. The column corresponding to the level of the predictor has the value 1, the other columns have the value 0. |
| **L1 & L2 penalty** | When an algorithm is fit to a dataset, a criterion is usually minimized, such as the sum of squared deviations. This is called a loss function. A penalty can be added to this loss function, influencing the fitted coefficients. This is called penalized or regularized fitting. A L1 (also called Lasso) penalty adds a term to the loss function based on the absolute values of predictor coefficients. This leads to models with less predictors. A L2 (also called Ridge) penalty adds a term to the loss function based on the square of the coefficients. This leads to smaller coefficients, which are also more evenly distributed between correlated predictors. |
| **SHAP values** | SHAP (SHapley Additive exPlanations) provide an explanation of how each predictor influences the model predictions for each individual prediction. They represent the marginal contribution of the predictor to the model. To calculate the total importance of a predictor, the mean of the absolute SHAP values is calculated. Higher values represent higher impact. |
| **Partial dependence plots** | These plots show the effect of changes in the values of a single predictor on the predicted outcome, marginalized over all other predictors. |
| **Leakage** | The use of data in a prediction which would not have been available at the time of prediction. This can cause overly optimistic estimates of predictive performance and suboptimal models. |
| **Simpson's paradox** | A statistical phenomenon where the association between a predictor variable and the outcome observed in the whole population could disappear or be reversed within subgroups. |

### Expert group meetings

Four meetings were held with an expert group of (wound care) nurses, researchers and a quality improvement nurse, discussing possible predictors, predictive performance, model development choices, and risks and barriers for implementation. Furthermore, group members were available for frequent unscheduled consultation by the model developers, who were data scientists.

### Data collection

Routinely collected electronic health records (EHR) data for each admission was obtained from the hospital’s data warehouse. The candidate predictors were chosen based on scientific evidence, on causal mechanisms (Coleman et al., 2014), predictive models (Wang et al., 2023), expert opinion from wound care nurses and nurse champions, and availability constraints.

Candidate predictors with near-zero variance were removed. Bivariate correlation coefficients and expert opinion were used to detect predictors with similar information and, when applicable, they were combined or adapted. The definition of the candidate predictors can be found in supplementary material table S1.

### Waterlow score

The Waterlow score was used in the hospital as the primary PU risk assessment tool and recorded by nurses. The Waterlow score includes 8 items: Body Mass Index (BMI), continence, skin status, mobility, sex, age, malnutrition, and specials risks including tissue malnutrition, neurological defects and major surgery or trauma (Waterlow, 2005). This results in a cumulative score ranging from 1 to 64 (highest risk). In this study it was used as a benchmark: the predictive performance for PU of the registered Waterlow scores was compared with the predictive performance of machine learning models.

### Prevalence screenings

Reliable PU prevalence screenings are performed three times per year in the selected hospital. These prevalence screenings are performed by wound care nurses and trained nurse champions, conducted according to the EPUAP statement on prevalence and incidence monitoring of PU occurrence (Defloor et al., 2005), which are considered the golden standard. The prevalence screenings were included as an additional data source for validation.

### Outcome measure for the model

PU at any point during the admission was used as a binary outcome. PU were registered at multiple places in the EHR: ICD-10 registration, structured registration in a nursing questionnaire, and free text from nurses’ reports. In the free text the presence of PU was written in several ways, including typos and spelling mistakes. To account for this, regular expressions were built and tested on the training dataset with the expert group to detect PU cases while preventing false hits.

The presence of PU is often underreported, with interobserver differences present (Crunden et al., 2022). To obtain an estimate of the amount of underreporting in the EHR, we compared the outcome measure from our training dataset to the PU prevalence screening. For the admissions with an observed PU during the prevalence screening, the percentage with a PU outcome documented in the training set was calculated.

### Dataset structure and split

The prediction model was intended to make daily predictions. The start of the nursing day shift (between 07:00 and 08:00 AM) was expected to be the primary time to consult the predictions, thus nightly predictions by the model were preferred. The nightly predictions could use the data up to midnight. The dataset structure was matched to these requirements, with a single row for each day during an admission. Separate models were developed for two distinct time windows: the first 72 hours of admission (≤72h), and the remainder of the admission (>72h). Since the dataset structure consisted of a separate row for each day during ad admission, often multiple rows were present for a single admission within the time windows. If a PU was registered, only rows in the admission prior to the first PU registration were retained. Subsequently, within the time windows a single row for each admission was obtained by random sampling.

Finally, the data was split into a training set (December 2021 – November 2022) and a validation set (January 2023 – June 2023).

### Data preprocessing

Daily values for each predictor were constructed. When multiple values for a single predictor were available on a day, aggregation was applied. The aggregation measure differed between predictors. For the vital parameters, the daily mean was calculated. For the medication administered, the daily sum was calculated. For all other predictors the most recent value from that day was used.

Data validation was applied to the retrieved data, transforming clearly implausible values caused by measurement error or data entry mistakes were transformed into missing values (Kahn et al., 2016). Numeric predictors were winsorized at the 0.005 and 0.995 quantiles, and standardized. Continuous predictors with a skewed distribution were transformed to a normal distribution (Yeo & Johnson, 2000), since skewed predictors can have a harmful effect on several types of machine learning models (Kuhn & Johnson, 2019). Categorical predictors were one-hot encoded (see Table 1).

Regression models estimate a single coefficient for continuous predictors, assuming a linear relationship between the predictor and the outcome. However, this assumption may not hold, and a non-linear relationship can be desirable. One way to achieve this is by dividing the predictor into several categories, however modelling non-linearity with a spline transformation is the preferred approach (Ma et al., 2023; Royston et al., 2006). When using a spline transformation, the relationship between the predictor and the outcome is divided into several windows, which are defined by so-called knots. Within each window, the relationship between the predictor and the outcome is modelled by a polynomial (Gauthier et al., 2020). This can make interpretation more difficult, however visualizing the relationship between the predictor and the outcome (‘partial dependency’) can aid in interpretation. The use of the spline transformation as a preprocessing step for a regression model was tested during comparison of the candidate models (see section ‘Candidate models’).

### Missing values

When data was not available on a daily basis the latest known value was filled forward for the admission. For instance, short-term repeated in-hospital albumin measurements are rarely performed, with a mean difference after 7 days with or without nutritional support of -0.72 g/L, which indicates that a value of at least one week before can still have a predictive value (Boesiger et al., 2023). Additionally, data from up to 7 days before the start of the admission could be used to forward-fill missing values, for instance questionnaires and lab results routinely collected before admission. The remaining missing values were imputed with the population mean, except for the expected remaining length of stay, for which the ward-specific median was imputed. Categorical and Boolean missing values were imputed with the most frequent category.

### Error analysis

After an initial round of dataset extraction and model development, an analysis was made of the prediction errors (averaged over the models) on the training set. Several cases with the highest amount of risk underestimation and overestimation were identified, with a follow-up of expert group consultation to identify the causes of the prediction errors. In some cases, relevant predictors were identified that were not available in the initial training set. When available in the EHR, they were added to the dataset.

### Candidate models

Eight models were trained on the candidate predictors and compared: logistic regression (LR) with either an L1 penalty (LR L1) or an L2 penalty (LR L2), logistic regression with a spline transformation and a L2 penalty (Spline LR L2), linear discriminant analysis (LDA), k-nearest neighbors (KNN), random forest (RF), extreme gradient boosting (XGB), and a single decision tree (Tree).

The area under the receiver operating characteristic curve (AUROC) was used to evaluate and compare model performance in class discrimination. The AUROC was estimated using repeated stratified cross-validation (see Table 1), consisting of 10 repeats of 5 folds, based on a patient level split. Also, for the subset of admission within the cross-validation fold with a recorded Waterlow score, both the model and the Waterlow AUROC were calculated on the fold’s validation data. Nested cross-validation (Cawley & Talbot, 2010) was applied to optimize model hyperparameters. Within each cross-validation fold, hyperparameters were optimized within the training data using 3-fold stratified cross-validation. Figure 1 illustrates the nested cross-validation within a single fold. A full list of the explored hyperparameters can be found in supplementary material table S2. Learning curves were made to visually assess whether the models were exposed to sufficient training samples (Figueroa et al., 2012). This was done by performing the cross-validation process with subsets of the dataset of different sizes (1000, 2000, 4000, 6000, 7548 (limit of >72h dataset), 11000, 15944 (limit of ≤72h dataset)) of the training data and visualizing the AUROC as a function of the size of the dataset.

**Figure 1.**
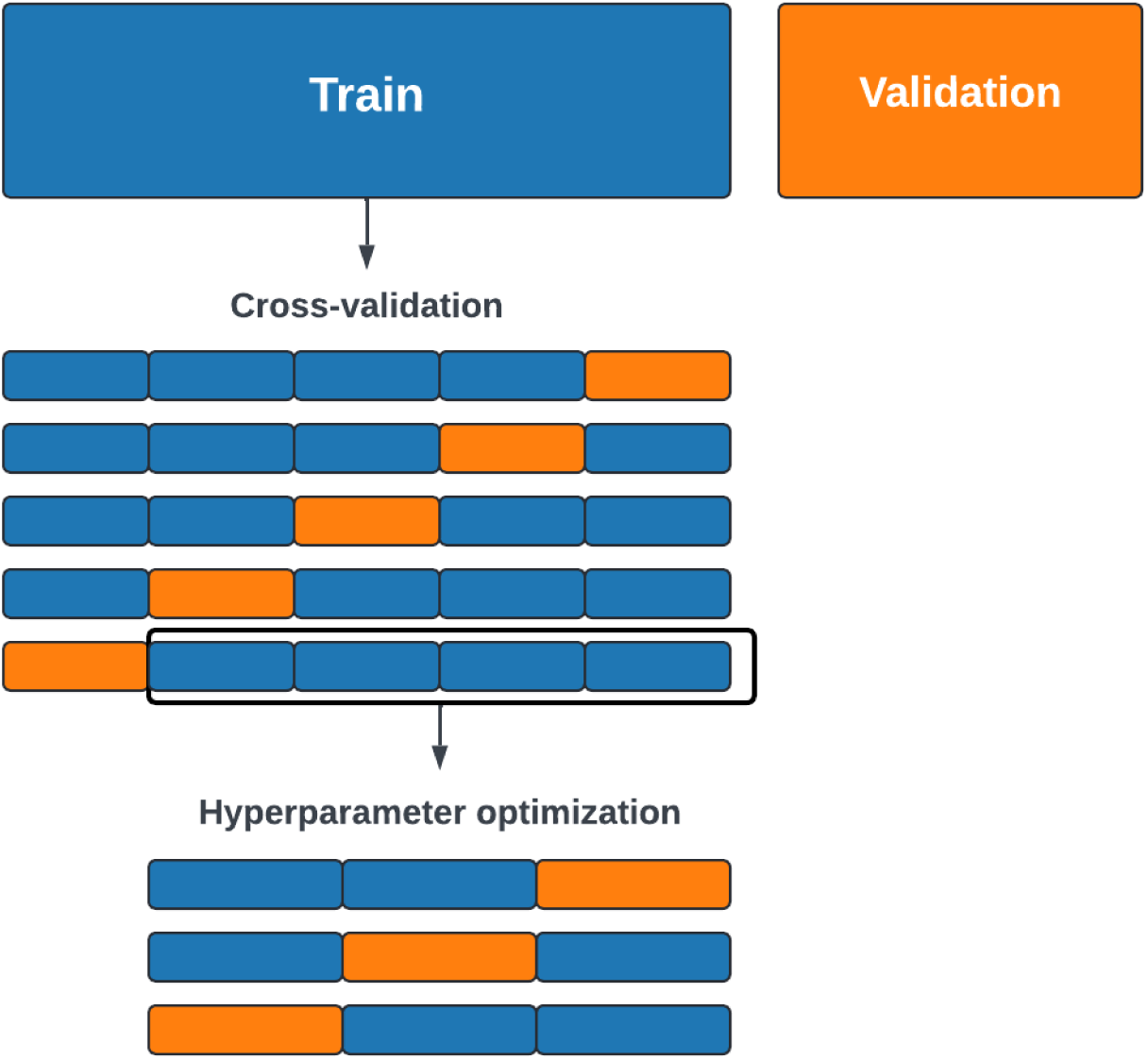
A single cross-validation iteration (out of 10) with nested hyperparameter optimization demonstrated for 1 of the 5 cross-validation folds.

The AUROC was also calculated on the prevalence screening datasets, with PU presence during the screening as the outcome measure. For each admission the first prediction available was used, which was made during the first night in the hospital. Admissions where the patient was screened before the first prediction were excluded, since no predictions were available. Admissions included in the prevalence screening were removed from the training set for the training of the candidate models, so the predictive performance of the candidate models on the prevalence screening dataset could be calculated in an out-of-sample manner.

### Selection of the final model and predictors

A final model was selected from the group of candidate models in accordance with the expert panel, based on the following considerations:

- Cross-validated predictive performance;

- Explainability;

- Expected robustness and longevity during clinical implementation.

Subsequently, predictor selection for the final model was performed. Predictors had to meet several criteria to be included:

- The predictor and its influence on the predictions should be explainable;

- The direction of the coefficient should be consistent with literature and expert group opinion, and the same for both models (≤72h & >72h);

- The magnitude of the coefficient should be large enough to make a noticeable impact on the predictions.

Since patients are clustered within wards, there are several subgroups present in the data. This introduces the risk of Simpson’s paradox (see Table 1) (Kievit et al., 2013). The possible occurrence of the paradox was checked by adding a fixed effect term for the ward and comparing the direction of the predictor coefficients for the model with the ward fixed effect to the final model without the fixed effect. After establishing the final predictor set, the final model was trained on the complete training set.

### Final model evaluation

The AUROC was calculated on the predictions of the final model for the validation datasets, and receiver operating characteristic (ROC) curves were made. Model calibration (see Table 1) was assessed using the calibration intercept and slope and visualized with a calibration curve (Van Calster et al., 2019). Clinical benefit was assessed using decision curve analysis (see Table 1) (Vickers & Elkin, 2006).

Model coefficients were reported, and predictor importance values were calculated using SHAP (SHapley Additive exPlanations) values (see Table 1) (Lundberg & Lee, 2017). The SHAP values were also used to create partial dependence plots (see Table 1). The distribution of the predictor values is also visualized in the partial dependence plots.

### Ethical considerations

The development of our prediction model followed existing laws, policies and in-company regulations. The key-principles for using AI in health from the WHO were considered during the development of the model (World Health Organization, 2021). The Medical Ethics Review Committee of the Erasmus University Medical Center assessed this study as not subject to the Dutch Medical Research Involving Human Subjects Act and confirmed that gathering informed consent for this study was not necessary (MEC-2023-0752).

## Results

### Dataset characteristics

A total of 19931 (≤72h) and 9435 (>72h) admissions were included in the training set. Since an admission can be present in both time windows, all 9435 admissions for the >72h window were also included in the ≤72h window. During 7.3% (≤72h) and 8.2% (>72h) of these admissions, the patient developed a PU. The validation set consisted of 11387 (≤72h) and 5201 (>72h) admissions. During 9% (≤72h) and 8.7% (>72h) of these admissions, the patient developed a PU. The descriptive statistics of the full candidate predictor set for ≤72h can be found in Table 2, including the percentage of missing values, which was over 50% for some predictors. The full description of the >72h set can be found in the supplementary material table S3.

**Table 2.** Descriptive statistics for the candidate predictor set, ≤72h time window. The column ‘Final model’ indicates whether the candidate predictor was included in the final model.

|  | Training (n = 19931) |  |  | Validation (n = 11387) |  |  | Final model |
| --- | --- | --- | --- | --- | --- | --- | --- |
|  | Missing | PU: False | PU: True | Missing | PU: False | PU: True |  |
| n (%) | 0.0% | 18474 (92.7) | 1457 (7.3) | 0.0% | 10365 (91.0) | 1022 (9.0) |  |
| Comorbidities: neuromuscular, n (%) | 0.0% | 598 (3.2) | 152 (10.4) | 0.0% | 310 (3.0) | 71 (6.9) | Yes |
| Comorbidities: peripheral vascular disease, n (%) | 0.0% | 276 (1.5) | 55 (3.8) | 0.0% | 155 (1.5) | 31 (3.0) | Yes |
| Comorbidities: liver, n (%) | 0.0% | 921 (5.0) | 111 (7.6) | 0.0% | 455 (4.4) | 55 (5.4) | Yes |
| Comorbidities: mobility, n (%) | 0.0% | 277 (1.5) | 64 (4.4) | 0.0% | 140 (1.4) | 34 (3.3) | Yes |
| Pressure ulcer during previous uptake, n (%) | 0.0% | 94 (0.5) | 85 (5.8) | 0.0% | 53 (0.5) | 25 (2.4) | Yes |
| Lab: albumin, mean (SD) | 76.2% | 31.7 (6.4) | 27.4 (7.2) | 76.2% | 31.3 (6.5) | 27.9 (6.9) | Yes |
| Lab: CRP, mean (SD) | 47.6% | 49.2 (75.9) | 83.5 (91.4) | 49.4% | 47.7 (71.2) | 72.6 (84.2) | Yes |
| Lab: RDW, mean (SD) | 22.3% | 14.3 (2.3) | 15.4 (2.7) | 22.2% | 14.3 (2.4) | 15.2 (2.6) | Yes |
| Lab: blood urea nitrogen, mean (SD) | 44.6% | 7.8 (5.7) | 9.6 (7.4) | 45.9% | 7.7 (5.6) | 10.0 (7.7) | Yes |
| Respiratory rate, mean (SD) | 10.3% | 15.5 (3.9) | 17.0 (4.7) | 9.5% | 15.8 (3.9) | 16.9 (4.6) | Yes |
| Heart rate, mean (SD) | 2.4% | 78.4 (14.4) | 85.1 (16.4) | 3.1% | 78.7 (14.3) | 83.4 (16.7) | Yes |
| Oxygen by mask, n (%) | 0.0% | 2022 (10.9) | 306 (21.0) | 0.0% | 1059 (10.2) | 190 (18.6) | Yes |
| Diastolic blood pressure, mean (SD) | 2.5% | 75.5 (11.1) | 72.6 (11.4) | 3.3% | 75.4 (10.9) | 73.4 (11.6) | Yes |
| Future surgery during stay, n (%) | 0.0% | 1215 (6.6) | 149 (10.2) | 0.0% | 589 (5.7) | 91 (8.9) | Yes |
| Surgery duration (minutes), mean (SD) | 57.8% | 167.9 (114.3) | 212.1 (151.6) | 56.6% | 166.7 (118.1) | 208.6 (140.1) | Yes |
| EMV score, mean (SD) | 71.6% | 14.7 (1.2) | 14.4 (1.6) | 70.5% | 14.7 (1.1) | 14.5 (1.4) | Yes |
| ADL Score, average of multiple questionnaires, mean (SD) | 27.8% | 0.5 (1.0) | 1.6 (1.4) | 24.8% | 0.4 (0.9) | 1.3 (1.4) | Yes |
| adapted MUST, mean (SD) | 19.4% | 0.3 (0.8) | 0.8 (1.3) | 16.4% | 0.4 (1.0) | 0.9 (1.3) | Yes |
| Freedom restricting interventions, n (%) | 0.0% | 167 (0.9) | 51 (3.5) | 0.0% | 77 (0.7) | 18 (1.8) | Yes |
| Medical devices score (0-4), mean (SD) | 0.0% | 0.2 (0.5) | 0.2 (0.6) | 0.0% | 0.2 (0.5) | 0.2 (0.5) | Yes |
| Incontinence, n (%) | 0.0% | 4066 (22.0) | 549 (37.7) | 0.0% | 2356 (22.7) | 329 (32.2) | Yes |
| ADL help required by nurses (0-1-2), mean (SD) | 0.0% | 0.3 (0.6) | 0.5 (0.7) | 0.0% | 0.3 (0.6) | 0.5 (0.7) | Yes |
| Non-elective admission, n (%) | 0.0% | 6769 (36.6) | 859 (59.0) | 0.0% | 3733 (36.0) | 608 (59.5) | Yes |
| Origin: from own home, n (%) | 0.0% | 16604 (89.9) | 1115 (76.5) | 0.0% | 9416 (90.8) | 803 (78.6) | Yes |
| Age, mean (SD) | 0.0% | 58.2 (16.7) | 64.1 (15.9) | 0.0% | 58.0 (16.7) | 63.4 (15.2) | Yes |
| Expected remaining length of stay (hours), mean (SD) | 17.5% | 72.5 (97.2) | 145.6 (139.2) | 15.2% | 68.9 (90.8) | 131.3 (124.0) | Yes |
| Comorbidities: kidney, n (%) | 0.0% | 1519 (8.2) | 190 (13.0) | 0.0% | 785 (7.6) | 103 (10.1) | No |
| Comorbidities: diabetes, n (%) | 0.0% | 2116 (11.5) | 298 (20.5) | 0.0% | 1120 (10.8) | 162 (15.9) | No |
| Comorbidities: cardiovascular, n (%) | 0.0% | 4520 (24.5) | 453 (31.1) | 0.0% | 2396 (23.1) | 304 (29.7) | No |
| Comorbidities: lung, n (%) | 0.0% | 2022 (10.9) | 231 (15.9) | 0.0% | 1151 (11.1) | 153 (15.0) | No |
| Lab: lactate, mean (SD) | 74.5% | 1.5 (1.0) | 1.6 (1.0) | 75.5% | 1.5 (1.1) | 1.6 (0.9) | No |
| Lab: hemoglobin, mean (SD) | 22.2% | 7.6 (1.4) | 7.0 (1.4) | 22.1% | 7.6 (1.4) | 7.2 (1.4) | No |
| Medication: vasopressors, n (%) | 0.0% | 2740 (14.8) | 116 (8.0) | 0.0% | 1739 (16.8) | 83 (8.1) | No |
| Medication: corticosteroids, n (%) | 0.0% | 4514 (24.4) | 270 (18.5) | 0.0% | 2807 (27.1) | 191 (18.7) | No |
| Medication: antihistamines, n (%) | 0.0% | 551 (3.0) | 38 (2.6) | 0.0% | 280 (2.7) | 19 (1.9) | No |
| Medication: chemotherapeutics, n (%) | 0.0% | 991 (5.4) | 58 (4.0) | 0.0% | 789 (7.6) | 45 (4.4) | No |
| Medication: anti-infectives, n (%) | 0.0% | 6810 (36.9) | 678 (46.5) | 0.0% | 3835 (37.0) | 443 (43.3) | No |
| O2 saturation, mean (SD) | 5.2% | 96.7 (1.8) | 96.2 (2.0) | 4.9% | 96.7 (1.9) | 96.3 (2.1) | No |
| Temperature, mean (SD) | 4.9% | 36.8 (0.5) | 36.9 (0.6) | 4.7% | 36.8 (0.5) | 36.9 (0.6) | No |
| Systolic blood pressure, mean (SD) | 2.5% | 129.7 (19.2) | 127.6 (20.2) | 3.3% | 129.4 (18.8) | 127.6 (20.8) | No |
| Average blood pressure, mean (SD) | 2.5% | 93.9 (12.8) | 91.4 (13.2) | 3.3% | 93.8 (12.5) | 91.9 (13.8) | No |
| Smoking, n (%) | 0.0% | 1929 (15.4) | 134 (13.0) | 0.0% | 1055 (14.7) | 96 (12.7) | No |
| Alcohol, n (%) | 0.0% | 4239 (35.9) | 279 (27.6) | 0.0% | 2503 (36.7) | 211 (29.0) | No |
| Drugs, n (%) | 0.0% | 381 (3.3) | 22 (2.2) | 0.0% | 230 (3.4) | 11 (1.6) | No |
| Gender: female, n (%) | 0.0% | 8158 (44.2) | 587 (40.3) | 0.0% | 4702 (45.4) | 455 (44.5) | No |
| BMI > 35, n (%) | 0.0% | 958 (6.4) | 63 (5.5) | 0.0% | 510 (5.9) | 51 (6.1) | No |

ADL: activities of daily living; BMI: body mass index; CRP: C-reactive protein; EMV: eye opening, best motor response, best verbal response; MUST: malnutrition universal screening tool; O2: oxygen; RDW: red blood cell distribution width. The definition of the candidate predictors can be found in supplementary material Table S1

### Outcome reporting in EHR

For all admissions with a PU during the prevalence screening, a PU was reported in the EHR for 78.6% of the training set and 74.6% of the validation set, indicating an underreporting of PU in the EHR in comparison with the prevalence screenings of 22.4% and 25.4% respectively.

### Predictor selection

After error analysis predictors indicating whether oxygen was administered by mask, freedom restricting interventions, a custom medical device score (based on the number and severity of medical devices in place), and neuromuscular disorders were added.

Furthermore, the detection of several comorbidities based on registered diagnoses was improved. In discussion with the expert group, predictors indicating physical therapy sessions and dietary consultations were not included in the candidate predictor set. Both predictors were deemed to bear more resemblance to a preventive measure than a risk factor. Table 2 lists the final predictor set.

### Comparing models

In general, predictive performance is shown to be slightly higher in the subset of admissions with a Waterlow score present, compared to admissions without. All machine learning models clearly outperform the Waterlow score. Figure 2 depicts the cross-validation AUROC results, with minimal differences between the models.

**Figure 2.**
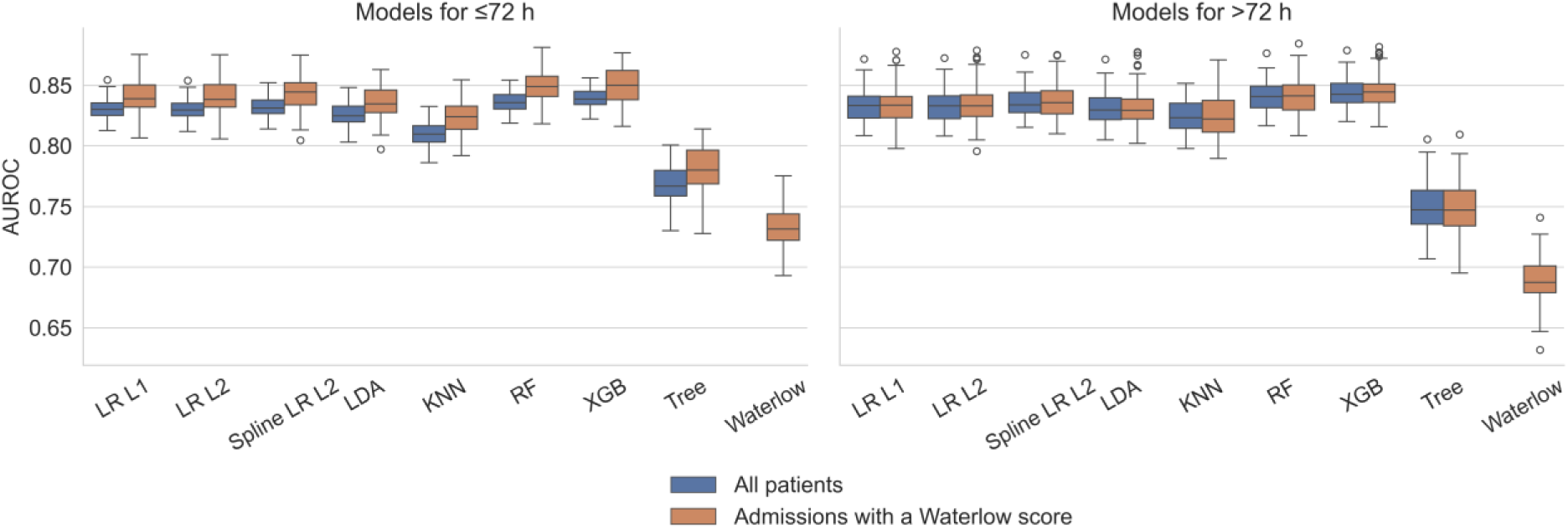
Cross-validated predictive performance for the candidate models and the Waterlow score. Predictive performance is measured by the area under the receiver operating characteristic curve (AUROC). The AUROC is calculated all admissions, and for the subset of admissions with a registered Waterlow score. LR: linear regression; LDA: linear discriminant analysis; KNN: k-nearest neighbors; RF: random forest; XGB: extreme gradient boosting.

The predictive performance on all prevalence screenings combined shows similar patterns, but with lower AUROC scores. Results for the predictive performance on the prevalence screenings can be found in supplementary material Table S4.

Learning curves (Figure 3) were made to visually assess whether the models were exposed to sufficient training samples. All models seemed to reach a plateau or be near a plateau in predictive performance, indicating the training set was of sufficient size. Since not all patients are admitted longer than 72 hours, the maximum number of samples differs between both graphs. Additionally, the maximum number of samples equals 80% of the training set size, since 5-fold cross-validation was used for assessing performance.

**Figure 3.**
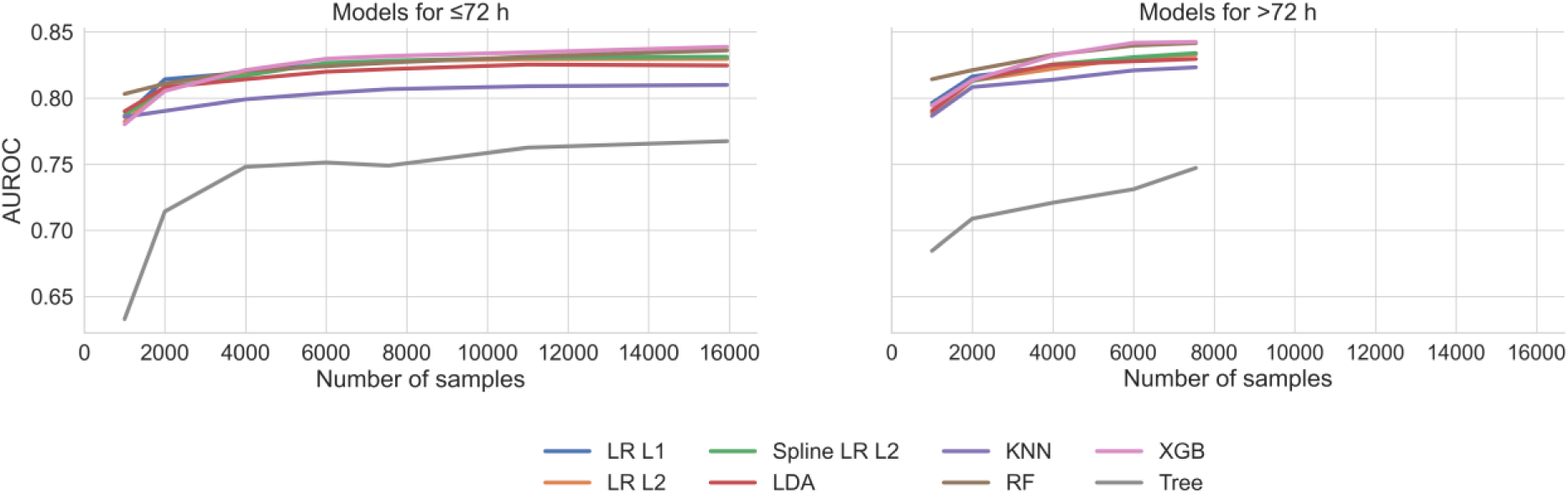
Learning curves of all the candidate models. The cross-validated predictive performance for the candidate models, as measured by the area under the receiver operating characteristic curve (AUROC), was calculated for subsets of the dataset of different sizes. LR: linear regression; LDA: linear discriminant analysis; KNN: k-nearest neighbors; RF: random forest; XGB: extreme gradient boosting.

### Final model

The final model was named DRAAI (Decubitus Risk Alert based on AI), which means ‘turn’ in Dutch and is a common PU preventive measure. Logistic regression with an L2 (ridge) penalty and spline transformation to a limited set of predictors was selected as the final model for both time windows and trained on the complete training set. Both the expert group and data-driven analysis indicated restricting all predictors to a linear coefficient was unrealistic. Since modelling non-linearity for all numeric predictors significantly increases the number of parameters to be estimated, it was only applied to a limited number of predictors with adequate data availability and for which the expert group and data-driven analysis strongly indicated non-linearity.

Table 3 summarizes the predictive performance of the final model on the validation datasets. Figure 4 displays the ROC curves.

**Figure 4.**
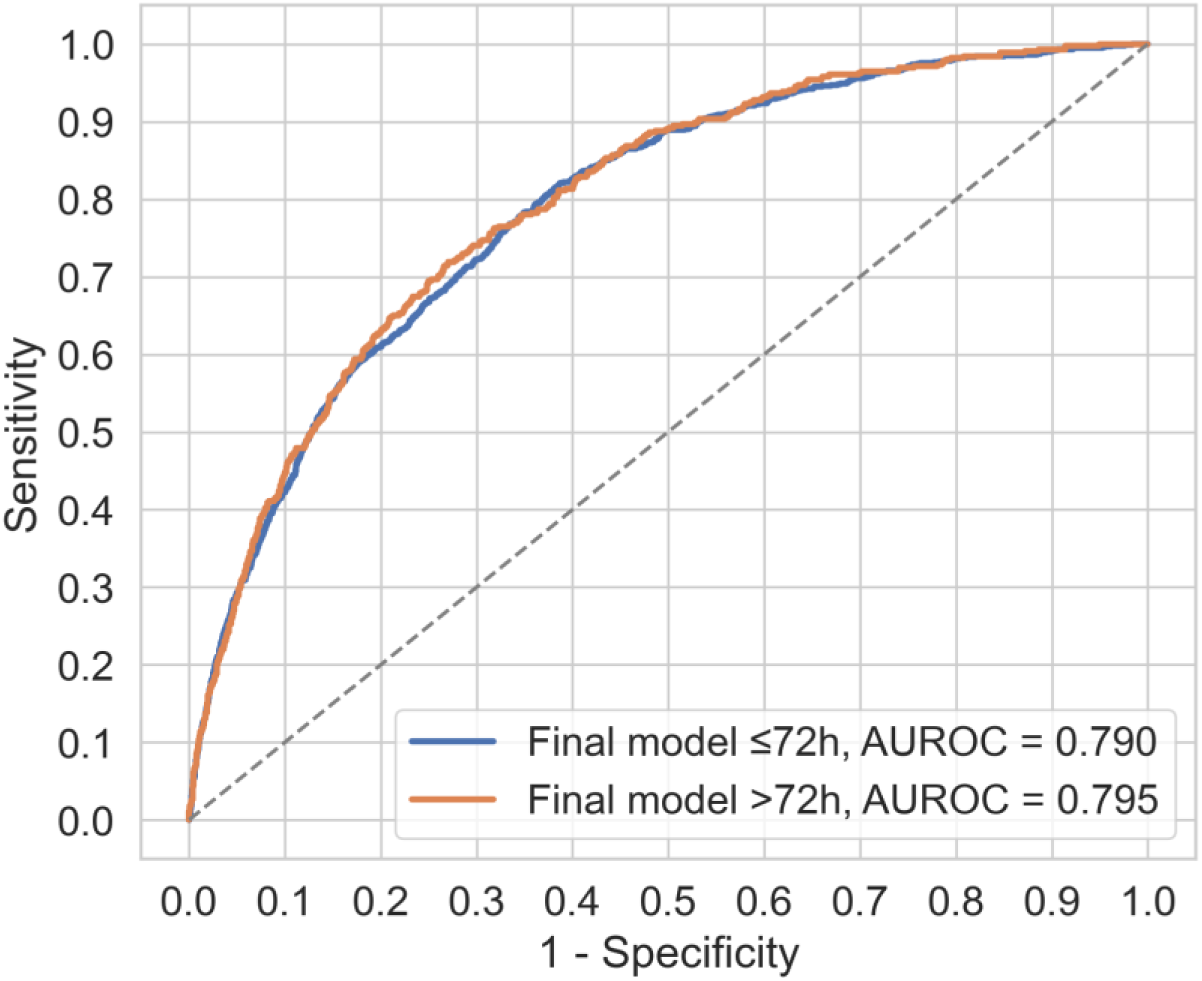
Receiver operating characteristic (ROC) curves for the final model on the validation dataset. AUROC: area under the receiver operating characteristic curve.

**Table 3.**
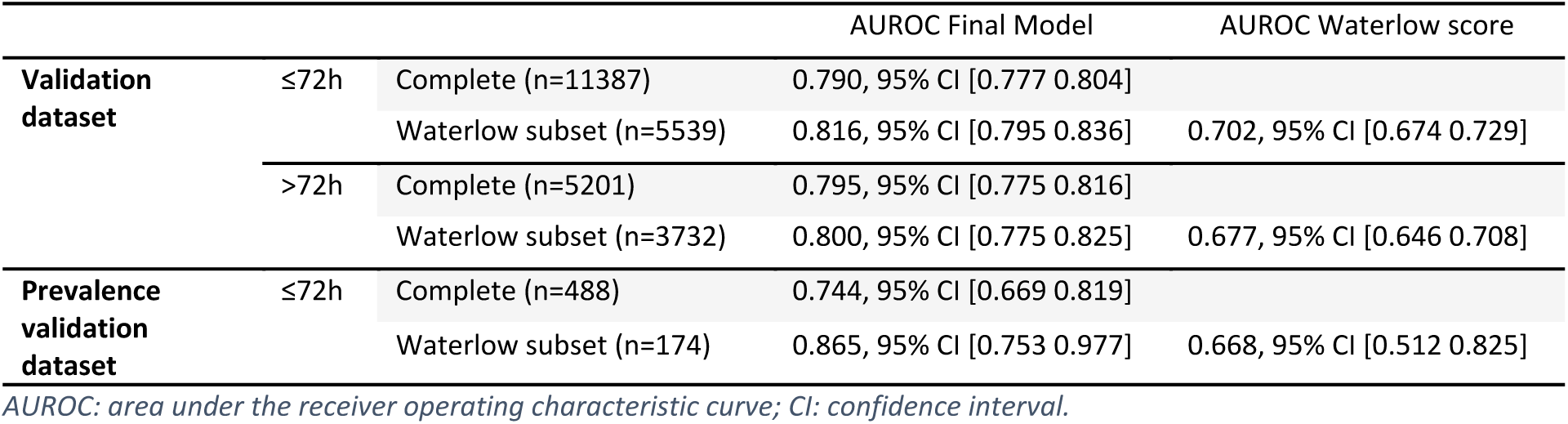
Predictive performance for the final model on the validation datasets.

The regression coefficients can be seen in Table 4. A spline transformation was applied to three predictor variables: ‘Age’, ‘Diastolic blood pressure’, and ‘Expected remaining length of stay’. As a result, for these predictors there is not a single regression coefficient, but a coefficient for each knot. Partial dependencies between these three predictors and the outcome for the ≤72h model can be found in Figure 5, for >72h they can be found in the supplementary material Figure S1. The top 10 predictors according to SHAP values are visualized in Figure 6, the SHAP values for all predictors can be found in the supplementary material Tables S5 and S6. No reversals of association direction between predictor and outcome due to Simpson’s paradox were detected (supplementary material Table S7).

**Figure 5.**
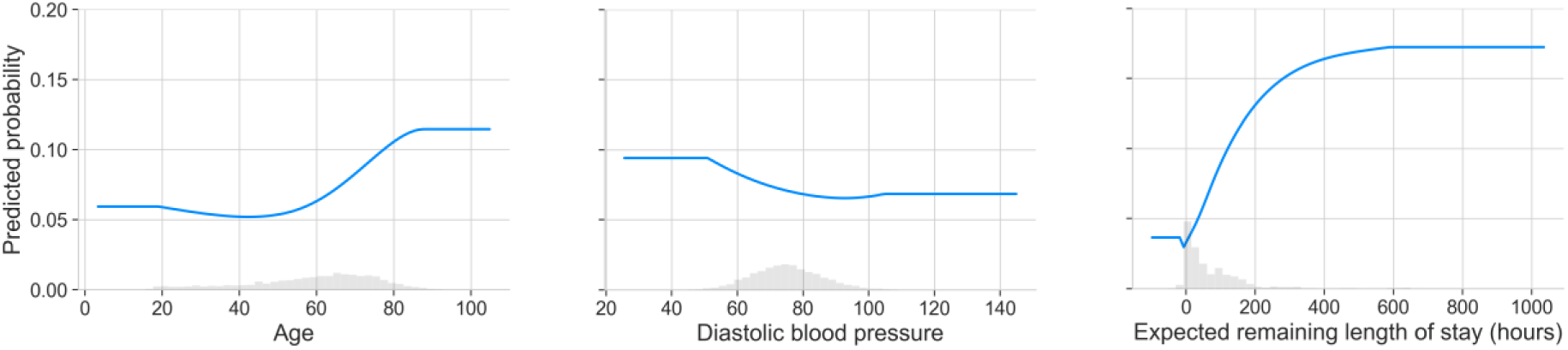
Partial dependence plots for the predictors with a spline transformation. These plots show the effect of changes in the values of a single predictor on the predicted outcome, marginalized over all other predictors.

**Figure 6.**
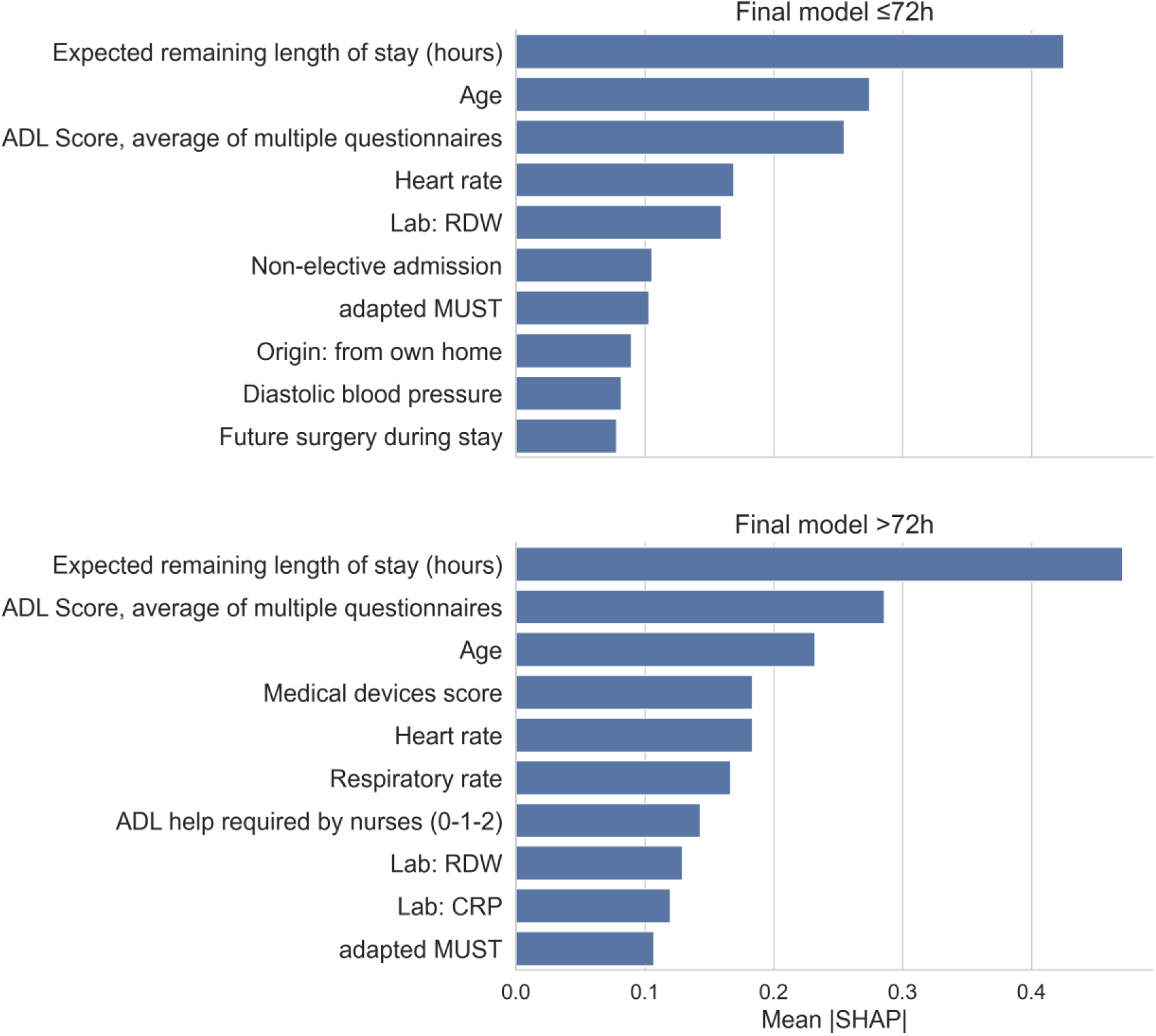
The 10 most important predictors for the final model as measured by the SHAP (SHapley Additive exPlanations) values. ADL: activities of daily living; CRP: C-reactive protein; MUST: malnutrition universal screening tool; RDW: red blood cell distribution width.

**Table 4.** Coefficients for the final model (logistic regression with a L2 penalty). A spline transformation was applied to three predictor variables: ‘Age’, ‘Diastolic blood pressure’, and ‘Expected remaining length of stay’. As a result, for these predictors there is not a single regression coefficient, but a coefficient for each knot.

| Predictor | Coefficient |  |
| --- | --- | --- |
|  | ≤72h | >72h |
| Intercept | -1.396 | -1.458 |
| ADL Score, average of multiple questionnaires | 0.406 | 0.362 |
| ADL help required by nurses (0-1-2) | 0.038 | 0.157 |
| Respiratory rate | 0.099 | 0.214 |
| Comorbidities: liver | 0.273 | 0.481 |
| Comorbidities: mobility | 0.504 | 0.172 |
| Comorbidities: neuromuscular | 0.700 | 0.441 |
| Comorbidities: peripheral vascular disease | 0.489 | 0.370 |
| EMV score | -0.155 | -0.236 |
| Freedom restricting interventions (Yes/No) | 0.335 | 0.630 |
| Heart rate | 0.212 | 0.227 |
| Incontinence | 0.201 | 0.093 |
| Age, spline knot 1 | 0.141 | -0.080 |
| Age, spline knot 2 | -0.739 | -0.735 |
| Age, spline knot 3 | -0.876 | -0.699 |
| Age, spline knot 4 | -0.145 | -0.204 |
| Age, spline knot 5 | 0.470 | 0.541 |
| Age, spline knot 6 | -0.246 | -0.281 |
| Lab: CRP | 0.153 | 0.183 |
| Lab: RDW | 0.240 | 0.163 |
| Lab: albumin | -0.153 | -0.244 |
| Lab: blood urea nitrogen | 0.037 | 0.116 |
| Medical devices score (0-4) | 0.031 | 0.245 |
| Surgery duration (minutes) | 0.192 | 0.068 |
| Origin: from own home | -0.541 | -0.353 |
| Oxygen by mask | 0.203 | 0.455 |
| Pressure ulcer during previous uptake | 1.857 | 0.657 |
| Non-elective admission | 0.225 | 0.175 |
| Adapted MUST | 0.190 | 0.161 |
| Diastolic blood pressure, spline knot 1 | 0.240 | 0.148 |
| Diastolic blood pressure, spline knot 2 | -0.091 | 0.243 |
| Diastolic blood pressure, spline knot 3 | -0.368 | -0.539 |
| Diastolic blood pressure, spline knot 4 | -0.555 | -0.544 |
| Diastolic blood pressure, spline knot 5 | -0.627 | -0.649 |
| Diastolic blood pressure, spline knot 6 | 0.003 | -0.117 |
| Future surgery during stay | 0.648 | 0.903 |
| Expected remaining length of stay (hours), spline knot 1 | 0.083 | -0.142 |
| Expected remaining length of stay (hours), spline knot 2 | -1.603 | -1.629 |
| Expected remaining length of stay (hours), spline knot 3 | -1.824 | -1.816 |
| Expected remaining length of stay (hours), spline knot 4 | 0.505 | 0.336 |
| Expected remaining length of stay (hours), spline knot 5 | 0.564 | 1.179 |

| Expected remaining length of stay (hours), spline knot 6 | 0.878 | 0.614 |
| --- | --- | --- |

ADL: activities of daily living; CRP: C-reactive protein; EMV: eye opening, best motor response, best verbal response; MUST: malnutrition universal screening tool; RDW: red blood cell distribution width.

Calibration analysis on the validation set resulted in intercepts of 0.02 (for both models) and slopes of 1.02 (≤72h) and 0.95 (>72h). Decision curves demonstrate the added value of using the models in clinical decision making over a broad range of plausible thresholds. The calibration plots (Figures S2 and S3) and decision curves (Figures S4 and S5) are visualized in the supplementary material.

## Discussion

We developed and validated a dynamic prediction model for daily pressure ulcer (PU) risk, meant for clinical application in a tertiary hospital. During the development process special care was given to considerations for implementation and user acceptance. A dataset with candidate predictors readily available in het EHR was used to compare several machine learning models, showing only minor differences in predictive performance across models. Logistic regression with an L2 penalty and a spline transformation applied to a limited set of predictors was chosen as the final model. Subsequently, the candidate predictor dataset was reduced to a final predictor set, and the final model was trained separately for two time windows during the admission: the first 72 hours of admission (≤72h), and the remainder (>72h). The AUROC for the final model was 0.790 (≤72h) and 0.795 (>72h) on the validation dataset. The model clearly outperformed the Waterlow score (0.816 vs. 0.702 (≤72h) and 0.800 vs. 0.677 (>72h)) for the subset of admissions with a registered Waterlow score.

Expected remaining length of stay, age and ADL score were the predictors with the strongest contribution.

Initially, multiple candidate machine learning models were compared, facilitating insight into a potential trade-off between predictive performance and interpretability (Markus et al., 2021). In contrast with some previous reports on PU prediction (Zhou et al., 2023), in our study little difference was found between models in terms of predictive performance. Given the small differences between models in predictive performance, the choice for a regression-based final model was based on three considerations. First, the additive nature of a regression model was preferred to interaction models (for example tree-based models) for explainability. Second, the final model should be able to sustain performance over time, while clinical care evolves. Regression models have been shown to suffer less performance degradation over time with non-stationary EHR data than interaction models, like tree-based ensemble models or neural networks (Jung & Shah, 2015; Nestor et al., 2019). Third, when the model should eventually be updated, there is more literature and guidance on updating regression models than algorithms like random forest or linear discriminant analysis (Meijerink et al., 2025).

Next, the candidate predictor set was reduced to a final predictor set. Because predictors can behave differently in different models, for example increasing predicted risk in a random forest while decreasing predicted risk in a logistic regression, the results of predictor selection methods depend on the chosen model (Pudjihartono et al., 2022). Thus, in the current study the manual predictor selection process was performed after choosing the final model. The approach to first select a model based on the full candidate predictor dataset, and then perform predictor selection, has been used before in PU prediction (Ladios-Martin et al., 2020). However, in the current study we extended the procedure by incorporating explainability and credibility as a criterion for predictor selection in addition to data driven selection methods to increase user acceptance of the model (Markus et al., 2021; Wang et al., 2018).

The model developed in this study was intended for clinical implementation in the same hospital setting as where it was developed. When validating the final model, the required type and extent of validation depend on the intended use of the prediction model and should match the population and setting intended for deployment (la Roi-Teeuw et al., 2024; Sperrin et al., 2022). Thus, in this study the dataset was split by calendar date, allowing for temporal validation (Ramspek et al., 2021; Steyerberg & Harrell, 2016). Model discriminative ability was measured with the AUROC and compared with the Waterlow score. To be able to compare threshold-dependent measures such as sensitivity and specificity to the Waterlow in a meaningful way, the threshold for the final model should be set so the Waterlow and the model designate an equal number of patients as ‘at risk’. However, the likelihood of this also being the optimal threshold for implementation in our clinical settings is small. Thus, in this study threshold dependent measures were not reported. However, they can be deduced from the ROC curve (Figure 4) for a range of thresholds.

Several potential modelling approaches were considered but not used in this study. First, with far fewer patients that developed PU during their admission than patients without, this is an imbalanced classification task, for which specialized methods like oversampling or undersampling exist. Previous research has shown these methods have no benefit in predictive performance for both PU specifically (Nakagami et al., 2021) and observational health data in general (van den Goorbergh et al., 2022; Yang et al., 2024). Thus, no class imbalance methods were used in this study. Second, most predictors in this study contained missing values, with some predictors having over 50% missing values. These missing values are often simply missing because some predictors such as lab values are not relevant for specific patients or that background characteristics were not documented. Advanced imputation procedures, like multiple imputation (Rubin, 1996) could perhaps prove beneficial to predictive performance. However, in discussions with the expert group and key users it became clear that a substantial influence of a missing value on the predicted risk for an individual patient might hinder explainability and user acceptance. Thus, simple methods of imputation were used to keep missing values uninformative. Last, hospital patients are clustered within wards, leading to a clustered data structure. Fixed effects models and random effects models are common statistical methods to account for data which is clustered within subgroups. Fixed effects models and random effects models include an intercept term for each subgroup in different ways, improving predictor coefficient estimates (Gardiner et al., 2009; McNeish & Kelley, 2019). In this study, no ward-specific intercept terms were used. The authors felt that since they would capture the influence of differing unmeasured variables, they would be susceptible to changes over time after clinical implementation of the prediction model. For example, personnel changes in a specific ward might change the priority of PU prevention, increasing or decreasing the prevalence.

Predictions from a model with an intercept term for that specific ward would then be systemically too low or too high. Not correcting for the clustering of patients within wards introduces the risk of Simpson’s paradox, but checks did not reveal issues in the current study.

### Strengths and limitations

A strength of our study is the frequency of predictions during a patient’s stay. Several studies have developed models with predictions on a single point in time, typically at the beginning of the stay (Cramer et al., 2019; Nakagami et al., 2021). However, during an admission, various risk factors might become relevant, some data might not be available at the beginning of the stay, and some data might become outdated. Accordingly, dynamic predictions during admission have been shown to outperform static predictions from the beginning of the stay (Shui et al., 2021). To accommodate dynamic predictions, (Shui et al., 2021) implemented four different models corresponding to different time windows during the admission: <24h, 24-48h, 48-72h, and >72h. However, using multiple models also increases day-to-day fluctuations in the risk scores, which then vary not only as a consequence of changes in predictor values, but also due to changes in model coefficients. Our expert group indicated that this might be a risk to user confidence and explainability. As a compromise, the current study used two time windows.

Another strength of this study is the careful consideration of the issue of data leakage when creating the dataset, which can occur when using predictor values which would have been unavailable at the time of prediction. Data leakage has been shown to be a widespread issue in machine learning and can cause overly optimistic performance estimates on retrospective datasets, with disappointing predictive performance when the model is implemented (Kapoor & Narayanan, 2023). When creating a historical data point used for machine learning, the predictor value known at that time should be used, instead of the latest value available. In this study database records of mutation were used to build retrospective datasets without leakage. For example, the expected discharge date can change several times during admission, however literature usually shows the use of a length of stay predictor (Wang et al., 2023), instead of the *expected remaining* length of stay used in this study.

A common limitation of practice is the documentation of PUs, which are often underreported. Also, healthcare providers are free to use free text registration, which limits structured reported data on the outcome (Chen et al., 2022; Crunden et al., 2022). In line with literature, PU were registered at multiple places in our EHR. Although this study supplemented structured registration with free-text detection, a comparison with the prevalence screenings revealed that not all PU were reported in the EHR. The inclusion of the prevalence screening datasets, which do not suffer this registrational issue, is a strength of this study. Next to indicating the amount of underreporting, they also provide an additional means of validation of the predictive performance.

Our study has a few limitations. The final model showed acceptable calibration, which could be improved by calibrating the predictions on the validation set. Ideally the models’ predictions would be calibrated to the risk of developing PU without any preventive measures. However, in clinical practice, the effect of historical policies on PU prevention will be included in any training dataset, lowering the outcome frequency and biasing calibration. Also, historical preventive interventions have not been included in the model. Furthermore, after successful implementation of the model in clinical practice, healthcare practitioners will act on the model predictions, lowering the outcome frequency even more. Thus, the precise meaning of the calibration of the predictions remains unclear, and the added value of the predictions lies primarily in the discriminative abilities.

### Implications for practice

After careful development of the prediction model, it is necessary to understand how to design successful implementation of the model in clinical practice. Acceptance and credibility of the model by professionals is key to healthcare providers’ adherence to the model’s predictions for preventive interventions, which then will lower the outcome frequency for patients identified as at risk for developing a PU. If later the model would be retrained on new data it would need to account for the effect of these model-based preventive interventions, otherwise the relationship between the predictors and the outcome will shrink towards zero and the model performance will be reduced (Khritankov, 2023; Lenert et al., 2019).

It is desirable to develop models which are transportable to other hospitals, in order to minimize resource waste. In practice, however, models developed in a specific setting (like our tertiary hospital) might be less performant in other settings, due to variations in factors such as patient population and measurement of predictors and outcomes, and due to change over time (Van Calster et al., 2023). Both the choice of predictors and the machine learning model may influence transportability (de Jong et al., 2021; Fehr et al., 2023). Several choices could be made when transporting, like retraining the model for different clinical settings, by training the model on combined data from multiple clinics (possibly through federated learning), or by combining multiple models (Reps et al., 2022). Although the model from this study was developed using input from both literature and the expert group, it should be considered to retrain or at least locally validate when transported to a different setting.

The future of predictive modelling in the field of predicting nurse-sensitive outcomes holds great promise, and the lessons learned from developing this prediction model for PU risk can be transferred to nurse sensitive outcomes such as falls, malnutrition, delirium or catheter related infections. The development process of such a model requires labor-intensive work from experts in multiple disciplines: both data scientist and experts in nursing. Expertise in several areas is required: policies and practices in the clinic and how they influence the data generated, knowledge of the EHR and data warehousing systems, how to generate datasets suitable for predictive modelling, statistical expertise in building robust and explainable machine learning models, and expertise in implementation in clinical routines. On top of that, monitoring and maintenance will be required after implementation, which should also be planned and budgeted. We recommend other developers and healthcare providers to closely work together with important stakeholders from the very beginning. Start with learning each other’s language, nurses have a completely different language than physicians or data scientists. Finally, in the implementation phase take into consideration that most of the time, the prediction model is not the only thing that needs to be implemented, the preventive measures are equally or even more important. Keep in mind that the ultimate goal is achieving a reduction in PU prevalence, one way or the other.

### Conclusions

We were able to develop and validate an explainable machine learning model that automatically provides dynamic PU risk predictions for hospitalized patients and clearly outperforms the Waterlow risk assessment score. Modelling choices were made in conjunction with an expert group of (wound care) nurses, increasing explainability and user acceptance. Furthermore, modelling choices were made to reduce the risk of performance degradation after clinical implementation.

## Data Availability

The patient level data is not publicly available.

## Acknowledgements

The authors acknowledge and thank all nurses who participated in this expert group for this study and especially acknowledge all nurses that have dedicated their time in preventing pressure ulcers. A special thanks to our project manager Daniëlle Janssen who managed to get all of us together, navigating us to all professionals concerning privacy, laws and regulations and her personal attention. We would also like to thank Joris Tukker who participated in the early phases of the project, and the team behind the Erasmus MC Data Platform (our data warehouse).

## Conflict of interest

None

## Funding sources

No external funding

## Notes

### Competing Interest Statement

The authors have declared no competing interest.

### Funding Statement

This study did not receive any external funding

### Author Declarations

The Medical Ethics Review Committee of the Erasmus University Medical Center waived ethical approval for this work. It assessed this study as not subject to the Dutch Medical Research Involving Human Subjects Act and confirmed that gathering informed consent for this study was not necessary (MEC-2023-0752).

