## Supplementary tables and figures for "Dynamic pressure ulcer risk predictions for hospitalized patients: development and validation of a machine learning model with an expert group of nurses"

FOR

**Affiliations**
[1] Department of Data & Analytics, Erasmus MC, Erasmus University Medical Centre, Rotterdam, The Netherlands
[2] Department of Internal Medicine, division of Nursing Science, Erasmus MC, Erasmus University Medical Centre, Rotterdam, The Netherlands
[3] Department of Quality and Patient Care, Erasmus MC, Erasmus University Medical Centre, Rotterdam, The Netherlands
[4] Department of Neonatal and Pediatric Intensive Care, division of Pediatric Intensive Care, Erasmus MC - Sophia Children's Hospital, Erasmus University Medical Centre, Rotterdam, The Netherlands

**Corresponding author**:
Steffen Greup
Erasmus MC, Erasmus University Medical Centre – Data & Analytics
P.O. Box 2040,
3000 CA Rotterdam, The Netherlands


#### Tables

***Table S1****. Definition of candidate predictors*

| Candidate predictor | Definition |
|---|---|
| Comorbidities: kidney | Anamnesis OR the following ICD10 codes: N12, N17 (valid for 100 days), N18, N19, N28.8 |
| Comorbidities: diabetes | Anamnesis OR the following ICD10 codes: E10, E11 |
| Comorbidities: neuromuscular | Anamnesis OR the following ICD10 codes: G20, G35, G63, G64, G70, G71, G80, G81, G82, G83, G95, I63 (valid for 730 days) |
| Comorbidities: cardiovascular | Anamnesis OR the following ICD10 codes: I10, I25, I48, I50, I69, I73, I79, M31.6, M31.7, M31.8, M31.9, R00.1, Z86.7 |
| Comorbidities: peripheral vascular disease | Anamnesis OR the following ICD10 codes: I73, I78, M31.8, M31.9 |
| Comorbidities: lung | Anamnesis OR the following ICD10 codes: J44 |
| Comorbidities: liver | Anamnesis OR the following ICD10 codes: K7 |
| Comorbidities: mobility | Anamnesis OR the following ICD10 codes: |

| | |
|---|---|
| | M86.65, M86.95, M86.97, S14, Z89.4, Z89.5, Z89.6, Z89.7, S12 (valid for 25 days), S72 (valid for 25 days), S82 (valid for 25 days), Z99 (valid for 25 days) |
| Pressure ulcer during previous uptake | ICD10 code L89 |
| Lab: albumin | |
| Lab: CRP | |
| Lab: lactate | |
| Lab: RDW | |
| Lab: hemoglobin | |
| Lab: blood urea nitrogen | |
| Medication: vasopressors | The following ATC codes:<br>C01CA04, C01CA03, C01CA24, H01BA01, C01CA07, C01CE02 |
| Medication: corticosteroids | The following ATC codes:<br>H02 |
| Medication: antihistaminica | The following ATC codes:<br>R06 |
| Medication: chemotherapeutics | The following ATC codes:<br>L01 |
| Medication: anti-infectives | The following ATC codes:<br>A02BD, A07AA, J01, J02, P |
| O2 saturation | |
| Temperature | |
| Respiratory rate | |
| Heart rate | |
| Oxygen by mask | True/False |
| Diastolic blood pressure | |
| Systolic blood pressure | |
| Average blood pressure | |
| Surgery duration (minutes) | |
| Future surgery during stay | True/False |
| EMV score | Overall score on the Glasgow Coma Scale, Eye opening, best Motor response, best Verbal response. |
| ADL Score, average of multiple questionnaires | Average of multiple questionnaires:<br>• The Katz-ADL overall score, except for the incontinence (which is a separate candidate predictor). 0-5 scale<br>• Anamnesis or transfer questions about the required help with mobilization, walking and personal care. Transformed to match the 0-5 scale. |
| adapted MUST | Overall risk score of the Malnutrition Universal Screening Tool. When the MUST was missing but the BMI was available, the MUST was imputed based on BMI:<br>BMI > 20: 0<br>BMI between 18.5 and 20: 1<br>BMI < 18.5: 3<br><br>In case of a BMI < 17, a point was added to the MUST score. |
| Freedom restricting interventions | True/False |
| Medical devices score (0-4) | Custom made 0-4 score. 1 point for Drains (True/False), 1 point for Catheters (True/False), 2 point for neck collars (True/False). |
| Incontinence | Based on Katz-ADL questionnaire item, and nursing orders in EHR. True/False |
| Smoking | Questionnaire result. True/False |
| Alcohol | Questionnaire result. True/False |
| Drugs | Questionnaire result. True/False |

| | |
|---|---|
| ADL help required by nurses (0-1-2) | Nursing orders in EHR for assisting the patient with ADL (True/False, 1 point) and mobilization (True/False, 1 point) |
| Non-elective admission | True/False |
| Origin: from own home | True/False |
| Gender: female | True/False |
| Age | |
| Expected remaining length of stay (hours) | The expected remaining length of stay at the time of prediction. |
| BMI > 35 | True/False |

***Table S2**. Definition of models and hyperparameters grids*

| | | |
|---|---|---|
| | Model | Logistic L1 |
| | | LogisticRegression(penalty = "l1", max_iter = 100000, solver = "liblinear") |
| tuning hyperparameter | C | [0.01, 0.1, 0.5, 1, 5] |
| | Model | Logistic L2 |
| | Definition | LogisticRegression(penalty = "l2", max_iter = 100000, solver = "liblinear) |
| tuning hyperparameter | C | [0.01, 0.1, 0.5, 1, 5] |
| | Model | Spline Logistic L2 |
| | Spline | SplineTransformer(degree=3, n_knots=4, knots = 'quantile') |
| | Definition | LogisticRegression(penalty = "l2", max_iter = 100000, solver = "liblinear") |
| tuning hyperparameter | C | [0.01, 0.1, 0.5, 1, 5] |
| | Model | LDA |
| | Definition | LinearDiscriminantAnalysis(solver = 'lsqr', shrinkage='auto') |
| | Model | KNN |
| | Definition | KNeighborsClassifier() |
| tuning hyperparameter | n_neighbors | [20, 40, 80, 160, 320, 640] |
| | Model | Random Forest |
| | Definition | RandomForestClassifier(n_estimators = 500) |
| tuning hyperparameter | max_depth | [3, 5, 8, 12] |
| | Model | XGBoost (Tree-based) |
| | Definition | XGBClassifier() |
| tuning hyperparameter | n_estimators | [40, 80, 160, 320] |
| tuning hyperparameter | learning_rate | [0.01, 0.1, 1] |
| tuning hyperparameter | max_depth | [3, 5, 8] |
| tuning hyperparameter | colsample_bylevel | [0.25, 0.5, 1] |
| | Model | Decision Tree |
| | Definition | DecisionTreeClassifier() |
| tuning hyperparameter | max_depth | [3, 5, 8, 12, 16] |

*Table S3*. *Descriptive statistics for the candidate predictor set, >72h time window*

| | Training (n = 9435) | | | Validation (n = 5201) | | |
|---|---|---|---|---|---|---|
| | Missing | PU: False | PU: True | Missing | PU: False | PU: True |
| n (%) | 0.0% | 8660 (91.8) | 775 (8.2) | 0.0% | 91.3) | 455 (8.7) |
| Comorbidities: neuromuscular, n (%) | 0.0% | 337 (3.9) | 52 (6.7) | 0.0% | 3.2) | 24 (5.3) |
| Comorbidities: peripheral vascular disease, n (%) | 0.0% | 144 (1.7) | 26 (3.4) | 0.0% | 1.7) | 10 (2.2) |
| Comorbidities: liver, n (%) | 0.0% | 526 (6.1) | 77 (9.9) | 0.0% | 5.6) | 33 (7.3) |
| Comorbidities: mobility, n (%) | 0.0% | 183 (2.1) | 26 (3.4) | 0.0% | 1.8) | 14 (3.1) |
| Pressure ulcer during previous uptake, n (%) | 0.0% | 47 (0.5) | 11 (1.4) | 0.0% | 0.7) | 6 (1.3) |
| Lab: albumin, mean (SD) | 49.7% | 28.3 (6.4) | 24.6 (6.4) | 50.4% | 28.3 (6.4) | 25.0 (6.8) |
| Lab: CRP, mean (SD) | 22.8% | 68.3 (77.3) | 95.0 (92.0) | 25.6% | 70.0 (75.9) | 93.4 (93.1) |
| Lab: RDW, mean (SD) | 4.7% | 14.7 (2.4) | 15.5 (2.6) | 6.6% | 14.6 (2.4) | 15.2 (2.5) |
| Lab: blood urea nitrogen, mean (SD) | 20.1% | 7.8 (6.0) | 9.9 (7.5) | 22.2% | 7.8 (6.0) | 10.1 (7.8) |
| Respiratory rate, mean (SD) | 5.6% | 15.3 (3.4) | 17.1 (4.5) | 6.7% | 15.7 (3.4) | 16.8 (4.6) |
| Heart rate, mean (SD) | 2.2% | 80.7 (13.8) | 86.8 (15.7) | 3.3% | 80.6 (13.6) | 83.8 (15.7) |
| Oxygen by mask, n (%) | 0.0% | 1036 (12.0) | 195 (25.2) | 0.0% | 536 (11.3) | 88 (19.3) |
| Diastolic blood pressure, mean (SD) | 2.5% | 74.9 (10.1) | 73.7 (11.4) | 3.6% | 75.0 (10.0) | 73.3 (10.9) |
| Future surgery during stay, n (%) | 0.0% | 335 (3.9) | 71 (9.2) | 0.0% | 128 (2.7) | 27 (5.9) |
| Surgery duration (minutes), mean (SD) | 47.8% | 227.8 (151.5) | 247.8 (187.6) | 48.4% | 227.1 (155.0) | 238.4 (171.4) |
| EMV score, mean (SD) | 52.5% | 14.8 (1.3) | 14.3 (1.6) | 51.3% | 14.8 (1.0) | 14.5 (1.5) |
| ADL Score, average of multiple questionnaires, mean (SD) | 10.8% | 0.8 (1.2) | 1.7 (1.4) | 7.7% | 0.7 (1.1) | 1.5 (1.4) |
| adapted MUST, mean (SD) | 5.8% | 0.4 (0.9) | 0.7 (1.2) | 4.2% | 0.5 (1.0) | 0.8 (1.3) |
| Freedom restricting interventions, n (%) | 0.0% | 185 (2.1) | 77 (9.9) | 0.0% | 79 (1.7) | 35 (7.7) |
| Medical devices score (0-4), mean (SD) | 0.0% | 0.3 (0.6) | 0.5 (0.7) | 0.0% | 0.3 (0.6) | 0.5 (0.7) |
| Incontinence, n (%) | 0.0% | 3029 (35.0) | 348 (44.9) | 0.0% | 1611 (33.9) | 179 (39.3) |
| ADL help required by nurses (0-1-2), mean (SD) | 0.0% | 0.6 (0.8) | 0.9 (0.8) | 0.0% | 0.6 (0.8) | 0.9 (0.8) |
| Non-elective admission, n (%) | 0.0% | 4297 (49.6) | 489 (63.1) | 0.0% | 2302 (48.5) | 272 (59.8) |
| Origin: from own home, n (%) | 0.0% | 7488 (86.5) | 604 (77.9) | 0.0% | 4187 (88.2) | 351 (77.1) |
| Age, mean (SD) | 0.0% | 58.5 (16.0) | 63.4 (15.5) | 0.0% | 58.5 (16.3) | 61.9 (16.1) |
| Expected remaining length of stay (hours), mean (SD) | 4.7% | 60.2 (79.7) | 123.2 (120.5) | 3.6% | 56.5 (74.4) | 109.8 (113.2) |
| Comorbidities: kidney, n (%) | 0.0% | 917 (10.6) | 100 (12.9) | 0.0% | 481 (10.1) | 42 (9.2) |
| Comorbidities: diabetes, n (%) | 0.0% | 1932 (22.3) | 277 (35.7) | 0.0% | 959 (20.2) | 138 (30.3) |
| Comorbidities: cardiovascular, n (%) | 0.0% | 2471 (28.5) | 249 (32.1) | 0.0% | 1294 (27.3) | 137 (30.1) |
| Comorbidities: lung, n (%) | 0.0% | 1926 (22.2) | 251 (32.4) | 0.0% | 1101 (23.2) | 133 (29.2) |
| Lab: lactate, mean (SD) | 50.9% | 1.3 (0.8) | 1.4 (0.7) | 53.1% | 1.3 (0.8) | 1.4 (0.9) |
| Lab: hemoglobin, mean (SD) | 4.7% | 7.0 (1.3) | 6.6 (1.3) | 6.5% | 7.0 (1.3) | 6.6 (1.3) |
| Medication: vasopressors, n (%) | 0.0% | 121 (1.4) | 25 (3.2) | 0.0% | 84 (1.8) | 27 (5.9) |
| Medication: corticosteroids, n (%) | 0.0% | 1990 (23.0) | 205 (26.5) | 0.0% | 1142 (24.1) | 120 (26.4) |
| Medication: antihistaminica, n (%) | 0.0% | 450 (5.2) | 30 (3.9) | 0.0% | 209 (4.4) | 18 (4.0) |
| Medication: chemotherapeutics, n (%) | 0.0% | 371 (4.3) | 34 (4.4) | 0.0% | 254 (5.4) | 16 (3.5) |

| | | | | | | |
|---|---|---|---|---|---|---|
| **Medication: anti-infectives, n (%)** | 0.0% | 3691 (42.6) | 482 (62.2) | 0.0% | 2129 (44.9) | 271 (59.6) |
| **O2 saturation, mean (SD)** | 2.3% | 96.5 (2.0) | 96.0 (2.0) | 3.3% | 96.5 (2.0) | 96.2 (1.9) |
| **Temperature, mean (SD)** | 1.5% | 36.8 (0.4) | 36.9 (0.6) | 2.5% | 36.8 (0.4) | 36.9 (0.5) |
| **Systolic blood pressure, mean (SD)** | 2.5% | 128.2 (18.0) | 129.0 (20.3) | 3.6% | 128.0 (17.8) | 128.1 (20.1) |
| **Average blood pressure, mean (SD)** | 2.5% | 92.7 (11.6) | 92.5 (13.3) | 3.7% | 92.7 (11.5) | 91.9 (13.1) |
| **Smoking, n (%)** | 19.4% | 959 (13.7) | 82 (13.7) | 16.9% | 524 (13.2) | 44 (12.1) |
| **Alcohol, n (%)** | 22.0% | 2310 (34.1) | 174 (29.9) | 19.6% | 1327 (34.6) | 92 (26.4) |
| **Drugs, n (%)** | 23.3% | 211 (3.2) | 15 (2.6) | 21.7% | 117 (3.1) | 3 (0.9) |
| **Gender: female, n (%)** | 0.0% | 3793 (43.8) | 339 (43.7) | 0.0% | 2132 (44.9) | 214 (47.0) |
| **BMI > 35, n (%)** | 5.9% | 552 (6.8) | 41 (5.8) | 4.4% | 260 (5.7) | 28 (6.7) |

***Table S4***. *AUROC on the prevalence screening training set*

| Model | AUROC Complete (n=707) | AUROC Waterlow subset (n=359) |
|---|---|---|
| Logistic L1 | 0.644, 95% CI [0.587 0.701] | 0.708, 95% CI [0.629 0.787] |
| Logistic L2 | 0.643, 95% CI [0.585 0.7] | 0.705, 95% CI [0.626 0.784] |
| Spline L2 | 0.637, 95% CI [0.579 0.695] | 0.68, 95% CI [0.599 0.76] |
| LDA | 0.643, 95% CI [0.586 0.701] | 0.705, 95% CI [0.627 0.782] |
| KNN | 0.646, 95% CI [0.588 0.703] | 0.7, 95% CI [0.617 0.783] |
| Random Forest | 0.657, 95% CI [0.599 0.714] | 0.713, 95% CI [0.631 0.796] |
| XGBoost | 0.657, 95% CI [0.599 0.715] | 0.701, 95% CI [0.619 0.783] |
| Tree | 0.593, 95% CI [0.531 0.654] | 0.65, 95% CI [0.556 0.745] |
| Waterlow score | | 0.625, 95% CI [0.528 0.721] |

***Table S5***. *Final model SHAP predictor importance values, ≤72h time window*

| Predictor | Mean \|SHAP\| |
|---|---|
| Expected remaining length of stay (hours) | 0.4251 |
| Age | 0.2744 |
| ADL Score, average of multiple questionnaires | 0.2546 |
| Heart rate | 0.1689 |
| Lab: RDW | 0.1593 |
| Non-elective admission | 0.1055 |
| adapted MUST | 0.1029 |
| Origin: from own home | 0.0895 |
| Diastolic blood pressure | 0.0814 |
| Future surgery during stay | 0.0779 |
| Lab: CRP | 0.0751 |
| Incontinence | 0.0746 |
| Respiratory rate | 0.0729 |
| Surgery duration (minutes) | 0.0689 |
| Comorbidities: neuromuscular | 0.0522 |
| Oxygen by mask | 0.0456 |
| Lab: albumin | 0.0302 |
| Comorbidities: liver | 0.0288 |
| ADL help required by nurses (0-1-2) | 0.026 |

| | |
|---|---|
| Medical devices score (0-4) | 0.0242 |
| EMV score | 0.0237 |
| Comorbidities: mobility | 0.0184 |
| Lab: blood urea nitrogen | 0.0175 |
| Pressure ulcer during previous uptake | 0.0167 |
| Comorbidities: peripheral vascular disease | 0.0128 |
| Freedom restricting interventions | 0.0102 |

***Table S6**. Final model SHAP predictor importance values, >72h time window*

| Predictor | Mean \|SHAP\| |
|---|---|
| Expected remaining length of stay (hours) | 0.4707 |
| ADL Score, average of multiple questionnaires | 0.2858 |
| Age | 0.2322 |
| Medical devices score (0-4) | 0.1834 |
| Heart rate | 0.1833 |
| Respiratory rate | 0.1666 |
| ADL help required by nurses (0-1-2) | 0.1429 |
| Lab: RDW | 0.1291 |
| Lab: CRP | 0.1197 |
| adapted MUST | 0.107 |
| Oxygen by mask | 0.1064 |
| Lab: albumin | 0.1054 |
| Origin: from own home | 0.0882 |
| Non-elective admission | 0.0877 |
| Diastolic blood pressure | 0.0876 |
| Lab: blood urea nitrogen | 0.0776 |
| Future surgery during stay | 0.0636 |
| EMV score | 0.0582 |
| Comorbidities: liver | 0.0559 |
| Comorbidities: neuromuscular | 0.0465 |
| Incontinence | 0.0457 |
| Surgery duration (minutes) | 0.034 |
| Freedom restricting interventions | 0.0294 |
| Pressure ulcer during previous uptake | 0.017 |
| Comorbidities: peripheral vascular disease | 0.0102 |
| Comorbidities: mobility | 0.0071 |

***Table S7**. Final model regression coefficients with or without Fixed Effects (FE) for the wards*

| Predictor | Coefficients ≤72h | | Coefficients >72h | |
|---|---|---|---|---|
| | Without FE | With FE | Without FE | With FE |
| ADL Score, average of multiple questionnaires | 0.406 | 0.399 | 0.362 | 0.34 |
| ADL help required by nurses (0-1-2) | 0.038 | 0.021 | 0.157 | 0.169 |
| Respiratory rate | 0.099 | 0.107 | 0.214 | 0.259 |
| Comorbidities: liver | 0.273 | 0.142 | 0.481 | 0.446 |

| | | | | |
|---|---|---|---|---|
| Comorbidities: mobility | 0.504 | 0.519 | 0.172 | 0.197 |
| Comorbidities: neuromuscular | 0.7 | 0.692 | 0.441 | 0.406 |
| Comorbidities: peripheral vascular disease | 0.489 | 0.514 | 0.37 | 0.482 |
| EMV score | -0.155 | -0.119 | -0.236 | -0.178 |
| Freedom restricting interventions | 0.335 | 0.399 | 0.63 | 0.584 |
| Heart rate | 0.212 | 0.217 | 0.227 | 0.243 |
| Incontinence | 0.201 | 0.182 | 0.093 | 0.084 |
| Age, spline knot 1 | 0.141 | 0.13 | -0.08 | -0.112 |
| Age, spline knot 2 | -0.739 | -0.763 | -0.735 | -0.739 |
| Age, spline knot 3 | -0.876 | -0.878 | -0.699 | -0.718 |
| Age, spline knot 4 | -0.145 | -0.129 | -0.204 | -0.173 |
| Age, spline knot 5 | 0.47 | 0.533 | 0.541 | 0.603 |
| Age, spline knot 6 | -0.246 | -0.251 | -0.281 | -0.265 |
| Lab: CRP | 0.153 | 0.142 | 0.183 | 0.212 |
| Lab: RDW | 0.24 | 0.243 | 0.163 | 0.174 |
| Lab: albumin | -0.153 | -0.139 | -0.244 | -0.257 |
| Lab: blood urea nitrogen | 0.037 | 0.053 | 0.116 | 0.127 |
| Medical devices score (0-4) | 0.031 | 0.011 | 0.245 | 0.238 |
| Surgery duration (minutes) | 0.192 | 0.177 | 0.068 | 0.096 |
| Origin: from own home | -0.541 | -0.534 | -0.353 | -0.316 |
| Oxygen by mask | 0.203 | 0.235 | 0.455 | 0.521 |
| Pressure ulcer during previous uptake | 1.857 | 1.876 | 0.657 | 0.695 |
| Non-elective admission | 0.225 | 0.177 | 0.175 | 0.146 |
| Adapted MUST | 0.19 | 0.187 | 0.161 | 0.176 |
| Diastolic blood pressure, spline knot 1 | 0.24 | 0.241 | 0.148 | 0.149 |
| Diastolic blood pressure, spline knot 2 | -0.091 | -0.068 | 0.243 | 0.287 |
| Diastolic blood pressure, spline knot 3 | -0.368 | -0.349 | -0.539 | -0.495 |
| Diastolic blood pressure, spline knot 4 | -0.555 | -0.541 | -0.544 | -0.514 |
| Diastolic blood pressure, spline knot 5 | -0.627 | -0.629 | -0.649 | -0.681 |
| Diastolic blood pressure, spline knot 6 | 0.003 | -0.012 | -0.117 | -0.149 |
| Future surgery during stay | 0.648 | 0.636 | 0.903 | 0.932 |
| Expected remaining length of stay (hours), spline knot 1 | 0.083 | 0.056 | -0.142 | -0.112 |
| Expected remaining length of stay (hours), spline knot 2 | -1.603 | -1.449 | -1.629 | -1.512 |
| Expected remaining length of stay (hours), spline knot 3 | -1.824 | -1.784 | -1.816 | -1.716 |
| Expected remaining length of stay (hours), spline knot 4 | 0.505 | 0.434 | 0.336 | 0.265 |
| Expected remaining length of stay (hours), spline knot 5 | 0.564 | 0.545 | 1.179 | 1.105 |
| Expected remaining length of stay (hours), spline knot 6 | 0.878 | 0.841 | 0.614 | 0.567 |

### Figures

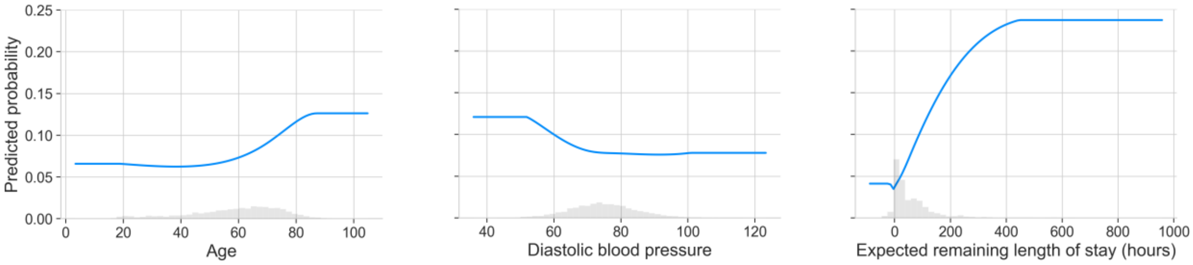

***Figure S1***. *Partial dependence plots for the predictors with a spline transformation.*

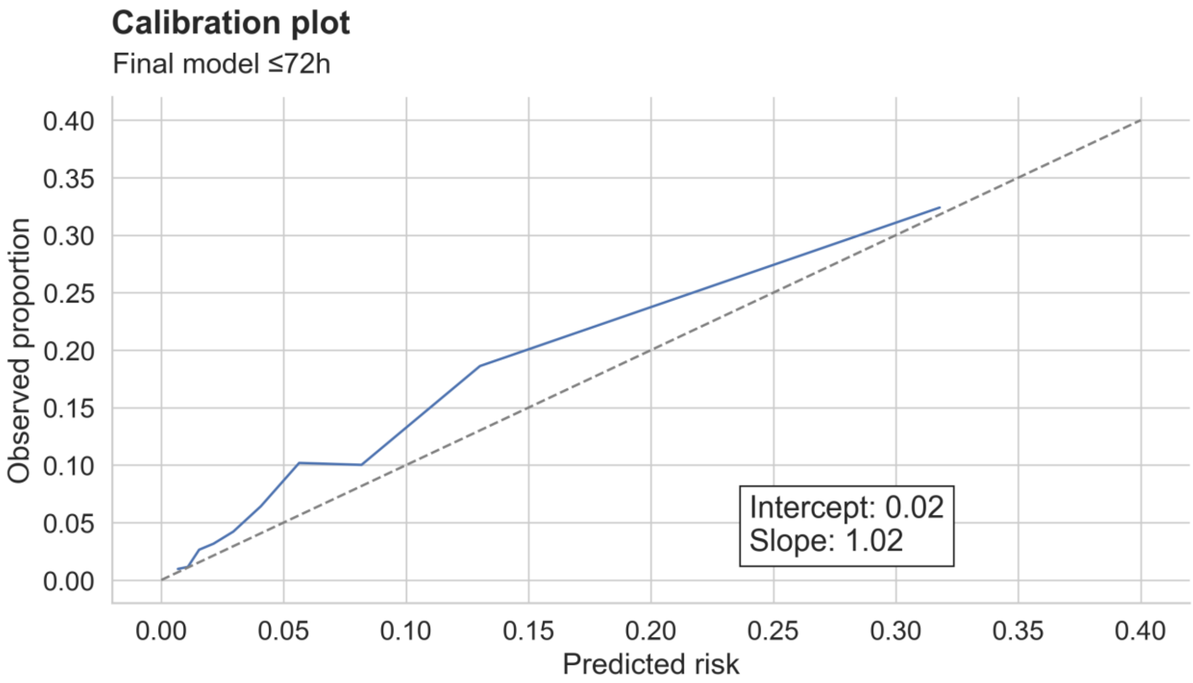

***Figure S2***. *Final model calibration plot, ≤72h time window*

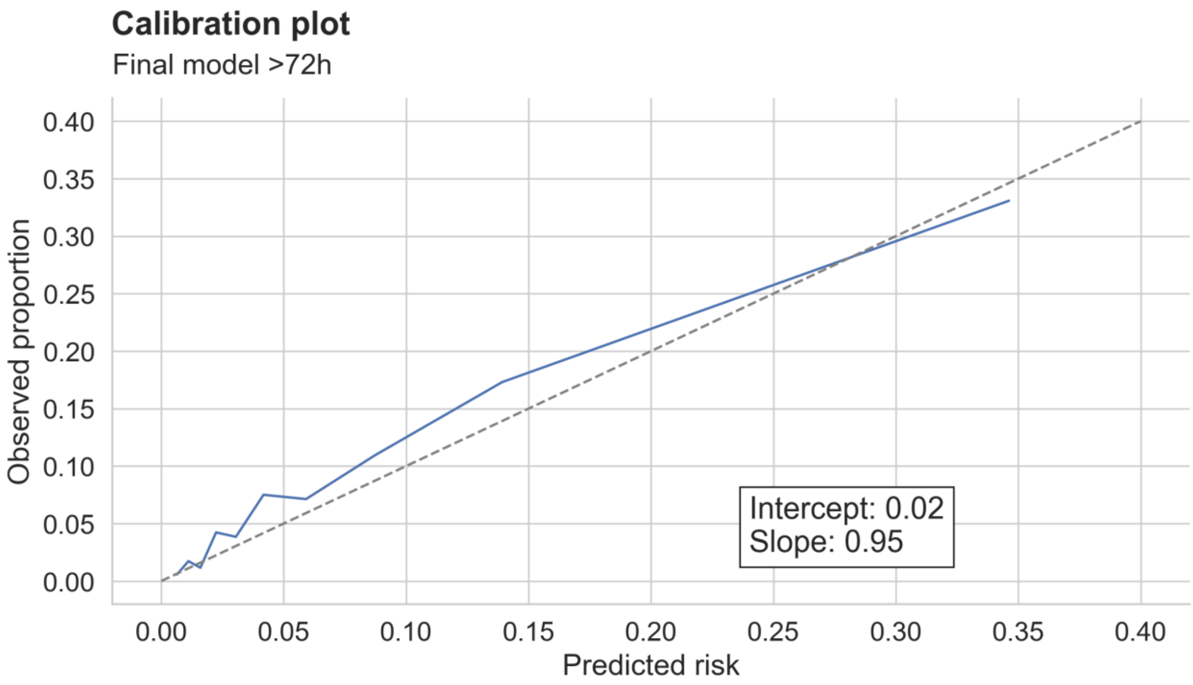

***Figure S3***. *Final model calibration plot, >72h time window*

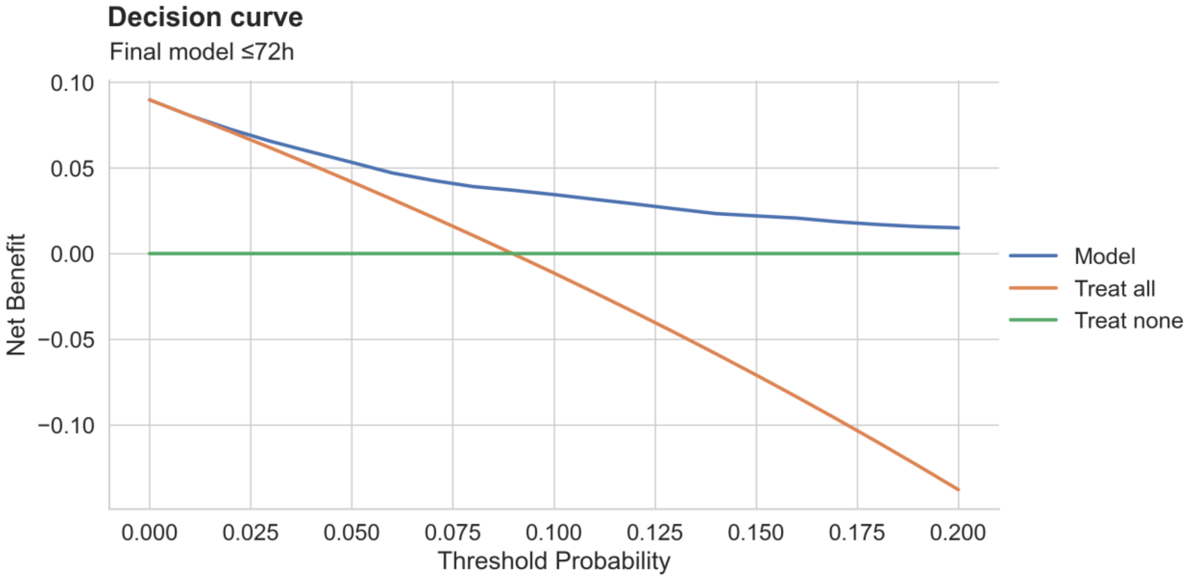

***Figure S4***. *Final model decision curve, ≤72h time window*

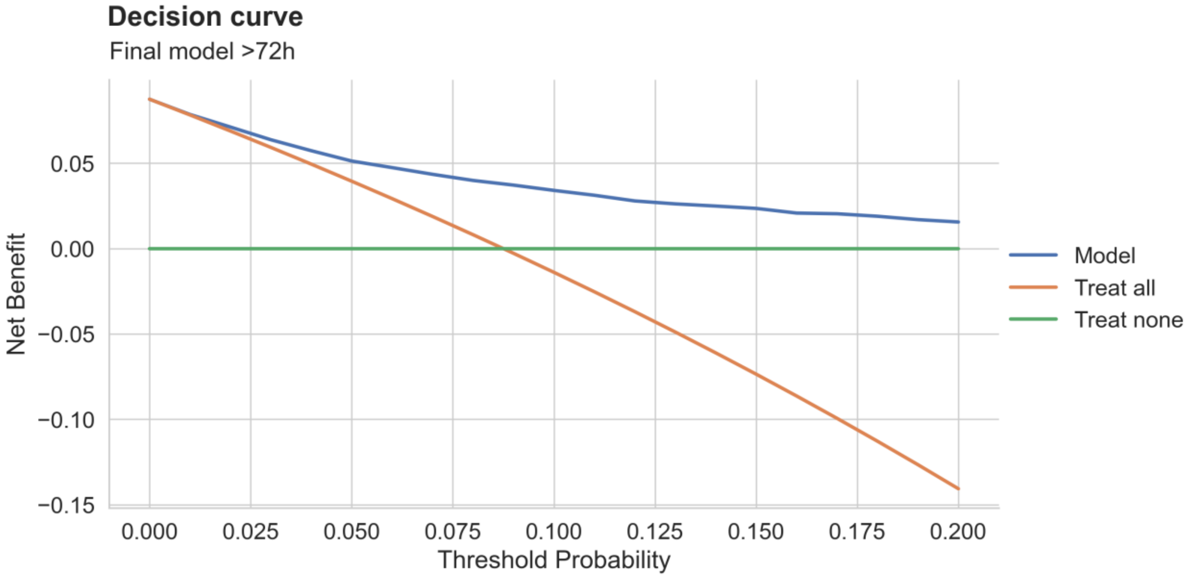

***Figure S5***. *Final model decision curve, >72h time window*
